# Large language model-based information extraction from free-text radiology reports: a scoping review protocol

**DOI:** 10.1101/2023.07.28.23292031

**Authors:** Daniel Reichenpfader, Henning Müller, Kerstin Denecke

## Abstract

**Introduction:** Radiological imaging is one of the most frequently performed diagnostic tests worldwide. The free text contained in radiology reports is currently only rarely utilized for secondary use, including research and predictive analysis. However, this data might be made available by means of information extraction (IE), based on natural language processing (NLP). Recently, a new approach to NLP, large language models (LLMs), has gained momentum and continues to improve performance. The objective of this scoping review is to show the state of research regarding IE from free-text radiology reports based on LLMs, to investigate applied methods, and to guide future research by showing open challenges and limitations of current approaches. To our knowledge, no systematic nor scoping review of IE of radiology reports, based on LLMs, has been conducted yet. Existing publications are outdated and do not comprise LLM-based models.

**Methods and analysis:** This protocol is designed based on the JBI manual for evidence synthesis, chapter 11.2: “Development of a scoping review protocol”. Inclusion criteria and a search strategy comprising four databases (PubMed, IEEE Xplore, Web of Science Core Collection, ACM Digital Library) are defined. Furthermore, we describe the screening process, data charting, analysis and presentation of extracted data.

**Ethics and dissemination:** This protocol describes the methodology of a scoping literature review and does not comprise research on or with humans, animals or their data. Therefore, no ethical approval is required. After the publication of this protocol and the conduct of the review, its results are going to be published in an open access journal dedicated to biomedical informatics/ digital health.

**Strengths and limitations of this study:**

- This scoping review protocol strictly adheres to standardized guidelines for scoping review conduction, including JBI Manual for Evidence Synthesis and the PRISMA-ScR guideline.
- The search strategy comprises four databases: PubMed, IEEE Xplore, Web of Science Core Collection, and ACM Digital Library.
- This scoping review will close the knowledge gap present in the field of information extraction from radiology reports caused by the recent rapid technical process.
- According to the nature of a scoping review, identified sources of evidence are not critically appraised.
- The results of the scoping review will serve as a basis for defining further research directions regarding information extraction from radiology reports.

## INTRODUCTION

Diagnostic tests like the many types of radiological imaging are the basis for decision-making in modern medicine (1): For example, 74.5% of Austrian women aged 50-69 have received bilateral mammography during the timeframe of two years according to the Austrian Health Interview Survey in 2019 (2). With breast cancer being the “second most common malignancy in the world” (3), mammography shows to reduce the risk of breast cancer mortality of women aged 50-69 with high certainty. This risk reduction is based on treatment decisions that are in turn based on radiology reports where experts describe the findings from the images. Traditionally, radiologists create semi-structured free-text radiology reports describing findings and their interpretation based on acquired images. Structured reporting, on the other hand, aims at improving clinical outcomes and standardization by providing frameworks for report layouts and contents. However, implementing structured reporting often requires changes to existing clinical processes. A consequent temporary increase in workload for radiologists makes it difficult to transfer structured reporting into clinical practice due to resistance among clinicians (4). Existing information could be made available by extracting clinically relevant information including its semantics and relations by applying natural language processing (NLP) methods. NLP is defined as the “tract of Artificial Intelligence and Linguistics, devoted to making computers understand the statements or words written in human languages” (5). Extracted information could be made available for secondary use, e.g., for prediction or research, based on methods related to information extraction (IE).

IE is a subfield within NLP to extract relevant information from text. Subtasks of IE include among others named entity recognition, relation extraction, and template filling. To solve these subtasks, different approaches might be applied: Basic approaches are based on heuristics. Machine learning-based approaches, on the contrary, include traditional methods (e.g., support vector machine, Naïve Bayes), or methods based on deep learning. Deep learning, in turn, comprises, among others, recurrent and convolutional neural networks as well as - most recently developed - large language models (LLMs) (6).

LLMs are «deep learning models with a huge number of parameters trained in an unsupervised way on large volumes of text» (7). We narrow this definition and only regard models with at least one million parameters as LLMs. Most of today’s models are based on the transformer architecture, which was first described in 2017 (8). Since then, new LLMs have been published on an ongoing basis, being trained on growing datasets and surpassing state-of-research performance regularly. Well-known models include BERT (2018, (9)), Megatron-ML (2019, (10)), GPT-3 (2020, (11)), GPT-4 and PaLM 2 (2023, (12,13)).

Regarding existing literature concerning IE from radiology reports, several reviews are available, although these sources either miss to include current developments or only focus on a specific aspect or clinical domain. Applying NLP to radiology reports for IE has already been focused on in two systematic reviews in 2016 (14) and 2021 (15). While the former is not freely available, the latter searches Google Scholar only and includes just one study based on LLMs. More recent reviews include a specific scoping review on the application of NLP to reports, specifically related to breast cancer (16) and a systematic review on the application of deep-learning-based NLP methods in radiology, although only including sources of evidence before 2019 (17). A search in PROSPERO, conducted on 30/05/2023, with the search query “Natural Language Processing AND radiology” yielded twelve results. Eleven results are not related to IE from radiology reports. One registered review describes named entity recognition and relation extraction in clinical documents using NLP. However, this review is neither focused on radiology reports nor LLM, and the search process was last updated on 07/07/2021, potentially missing many of recently published articles regarding the application of LLMs (18). Therefore, as LLMs have only recently gained momentum, a research gap exists and there is no overview of LLM-based approaches to IE from radiology reports available.

As compared to a systematic review, a scoping review usually does not include a critical appraisal of the identified sources of evidence. On the other hand, conducting a scoping review takes fewer resources to perform and is therefore especially suitable for the dynamically changing research area focused on IE from radiology reports. With this protocol for a scoping review, we therefore intend to fill the identified research gap and answer the following research question:

### What is the state of research regarding information extraction from free-text radiology reports based on Large Language Models?

Specifically, we are interested in the sub-questions that arise from the posed research question, see Table 1.

**Table 1:** Research sub-questions to be answered based on the scoping review.

|  |  |
| --- | --- |
| RQ.01 – Performance | What is the performance of LLMs for information extraction from radiology reports? |
| RQ.02 – Training and Modelling | Which models are used and how is the pre-training and fine-tuning process designed? |
| RQ.03 – Use cases | Which modalities and anatomical regions do the analyzed reports correspond to? |
| RQ.04 – Data and annotation | How much data was used to train the model, how was the annotation process designed and is the data publicly available? |
| RQ.05 – Challenges | What are open challenges and common limitations of existing approaches? |

The objective of this scoping review protocol is to answer the above-mentioned aspects, give an overview of recent developments, and guide future research by showing open challenges and limitations of current approaches.

## METHODS AND ANALYSIS

The scoping review will adhere to the JBI Manual for Evidence Synthesis, chapter 11: Scoping reviews (19). This manual in turn complies with the specifications of the PRISMA Extension for Scoping Reviews (PRISMA-ScR), which provides a guideline on the design and methodology of a scoping review (20).

This protocol is designed specifically based on chapter 11.2 of the JBI manual: “Development of a scoping review protocol”. The manual defines sections and their contents to be included in the protocol, comprising inclusion criteria, search strategy, source of evidence selection, data extraction, analysis of the evidence, and presentation of results. These aspects are described in the following chapters.

### Inclusion criteria

In Table 2, we describe the criteria to be applied in selecting sources of evidence (SOE). Focus was put on aligning these criteria with the title as well as the research question and sub-questions of the scoping review.

**Table 2:** Inclusion criteria.

|  |  |
| --- | --- |
| C.01 | The full-text SOE is retrievable. |
| C.02 | The SOE was published after 31/12/2017. |
| C.03 | The SOE is published in a peer-reviewed journal or conference proceeding. |
| C.04 | The SOE describes original research, excluding reviews, comments, patents, and white papers. |
| C.05 | The SOE describes the application of NLP methods for the purpose of IE from free-text radiology reports. |
| C.06 | The described approach is LLM-based (defined as deep learning models with more than one million parameters, trained on unlabeled text data). |

### Search strategy

The chosen search strategy comprises three steps: First, a limited search of at least two databases (PubMed, Google Scholar) is used to obtain a list of relevant index terms and keywords, see Table 3. Next, based on this list of terms, a comprehensive and systematic search query is developed iteratively.

**Table 3:** Primary search terms.

|  |  |
| --- | --- |
| PubMed | (information extraction) AND (radiolog*) AND (report*) AND (large language model) |
| Google Scholar | “information extraction radiology reports large language model” |

We include four databases to be searched using the developed query: PubMed, IEEE Xplore, Web of Science Core Collection, and ACM Digital Library. The primary search query will be developed for usage with PubMed and then translated to be used for the other three databases, where possible automatically (21). Each of the four search strings, including the number of retrieved records, date coverage, and date of search, will be documented using a standardized template provided by Karolinska Institutet (22).

As a third and last step, after the selection process, reference lists of studies that are included in the review are searched for additional sources of evidence (“forward-search”). This process might be supported by automation tools.

### Source of evidence selection

The SOE selection process will be conducted by two reviewers individually. The review process is performed and managed using the software platform Rayyan (23). Before screening, duplicate records are removed semi-automatically (manual check of automatically identified duplicates) and a pilot testing procedure is carried out to ensure agreement of both reviewers on inclusion criteria: A random sample of 25 SOE entries is selected and assessed by both reviewers. Then, decisions are compared. In case of any differences, inclusion criteria are clarified and/or adapted. Screening is started only when an agreement of >75% is achieved – otherwise, additional batches of ten SOE entries are assessed similarly until the specified level of agreement is reached.

Next, all records, consisting of titles and abstracts, are screened by both reviewers and included if they fulfill all inclusion criteria. After completion, disagreements are solved by the decision of a third reviewer. Then, full-text retrieval is performed for all included records. Records that cannot be retrieved are excluded. Retrieved full texts are assessed for eligibility: Sources that do not comply with all defined inclusion criteria are excluded. Last, a forward search is performed using reference lists of remaining sources of evidence. See Figure 1 for an illustration of the described process.

**Figure 1:**
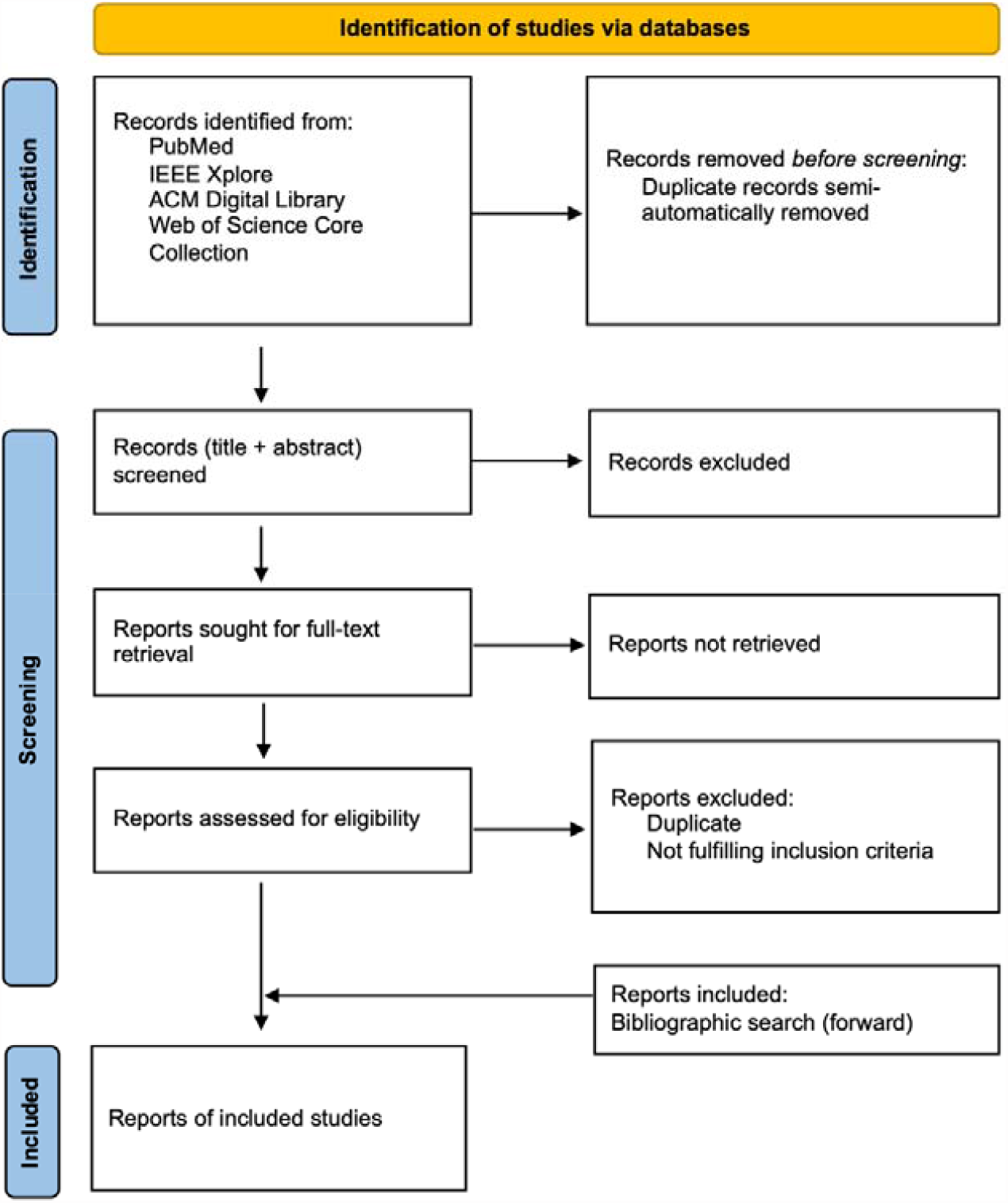
Source of evidence selection process

### Data extraction

As a next step, key information is extracted from the final set of included studies. A charting table was created based on the JBI manual, Appendix 11.1, and adapted as well as augmented in accordance with the research question and sub-questions this scoping review addresses, see Table 4 (19). Before extraction, a pilot test is conducted first to ensure the validity of the data charting table: Two sources of evidence are extracted by two reviewers, and results as well as possible adaptions of the charting table are discussed and agreed on. Upon agreement, data extraction is performed by one reviewer.

**Table 4:** Data charting table.

|  |
| --- |
| **Scoping review details** |
| Scoping review title |
| Review objective |
| Review question and sub-questions |
| **Evidence source: Details and Characteristics** |
| Citation details (e.g. author/s, date, title, journal, volume, issue, pages) |
| Origin/country of origin |
| **Details extracted from source of evidence (acc. to sub-questions)** |
| **Extracted information** |
| Information model (description of entities and/or relations) |
| Information model development process |
| Structuring of results (e.g. mapping to ontology) |
| **Model** |
| Model design |
| Pre-training and further pre-training process |
| Fine-tuning process |
| Described performance measures |
| Baseline |
| **Data set** |
| Amount |
| Split training/test/validation |
| Availability |
| Modality |
| Anatomical region |
| Origin |
| Language |
| **Annotation process** |
| Process description |
| Approach (automated, semi-automated, manual, mixed) |
| Number of annotators |
| Annotation guideline |
| Inter-Annotator Agreement |
| Tools used |
| **Data availability (source code)** |
| **Open challenges** |
| **Limitations** |

### Analysis of the evidence and presentation of results

Analysis of evidence is limited to descriptive mapping and does not include synthesis or critical appraisal. Aspects described in the data charting table are described by frequency counts where possible. These frequencies provide the basis to answer the research sub-questions described in Table 1. Results are presented using either tables, lists, crosstabulations, bar charts, pie charts or other diagram types. Diagrams and tables are accompanied by descriptive texts.

## Data Availability

All data produced in the present work are contained in the manuscript

## Ethics and dissemination

This scoping review protocol does not include any research with or related to humans, animals or their data, hence no ethical approval is sought for. After the publication of the protocol, the scoping review itself is carried out. Its results are then published in an open access journal dedicated to the field of biomedical informatics.

## Author’s contributions

Daniel Reichenpfader: Conceptualization, Methodology, Writing – Original Draft Kerstin Denecke: Writing - Review & Editing

Henning Müller: Supervision

